# Development and validation of artificial intelligence-based algorithms for predicting the segments debulked by rotational atherectomy using intravascular ultrasound

**DOI:** 10.1101/2023.11.07.23298239

**Authors:** Kenta Hashimoto, Kenichi Fujii, Daiju Ueda, Akinori Sumiyoshi, Katsuyuki Hasegawa, Rei Fukuhara, Munemitsu Otagaki, Atsunori Okamura, Wataru Yamamoto, Naoki Kawano, Akira Yamamoto, Yukio Miki, Iichiro Shiojima

## Abstract

**Background:** Although rotation atherectomy (RA) is a useful technique for severely calcified lesions, patients undergoing RA show a greater incidence of catastrophic complications, such as coronary perforation. Therefore, prior to the RA procedure, it is important to predict which regions of the coronary plaque will be debulked by RA.

**Objectives:** We develop and evaluate an artificial intelligence–based algorithm that uses pre-RA intravascular ultrasound (IVUS) images to automatically predict regions debulked by RA

**Methods:** A total of 2106 IVUS cross-sections from 60 patients with de novo severely calcified coronary lesions who underwent IVUS-guided RA were consecutively collected. The two identical IVUS images of pre-and post-RA were merged, and the orientations of the debulked segments identified in the merged images are marked on the outer circle of each IVUS image. The artificial intelligence model was developed based on ResNet (deep residual learning for image recognition). The architecture connected 36 fully connected layers, each corresponding to one of the 36 orientations segmented every 10°, to a single feature extractor.

**Results:** In each cross-sectional analysis, our artificial intelligence model achieved an average sensitivity, specificity, positive predictive value, negative predictive value, and accuracy of 81%, 72%, 46%, 90%, and 75%, respectively.

**Conclusions:** The artificial intelligence–based algorithm can use information from pre-RA IVUS images to accurately predict regions debulked by RA. The proposed method will assist interventional cardiologists in determining the treatment strategies for severely calcified coronary lesions.

## Introduction

Percutaneous coronary intervention (PCI) in patients with severe coronary artery calcification poses technical challenges due to impaired device crossing(1), delamination of drugs and polymers from stents(2), altered elution kinetics and drug delivery(3), and impaired stent apposition and expansion(4). Rotational atherectomy (RA) is used to modify lesions to allow optimal stent deployment and expansion and to improve clinical outcomes in patients with severely calcified coronary lesions(5). Recently published North American and European expert reviews and the Japanese expert consensus on RA showcase its utility and provide a clinical standard for RA operators(6–8).

Although RA is a useful technique for severely calcified lesions, patients undergoing RA show a significantly greater incidence of catastrophic complications (e.g., slow flow, complex dissection, and coronary perforation) than that in patients undergoing balloon-based procedures(9,10). Therefore, one expert consensus document recommends using intravascular imaging devices prior to RA to predict the site of RA debulking using a guidewire and catheter bias(8). However, the agreement rate between the guidewire position and the debulked region on intravascular imaging, such as intravascular ultrasound (IVUS), is not high(11,12); therefore, predicting the debulking site with intravascular imaging devices remains a challenge, especially for less experienced interventional cardiologists.

Deep learning techniques that implement deep neural networks can extract features from target data without preconceptions and are, therefore, particularly useful for detecting and classifying objects with complicated or unknown features(13). Consequently, deep learning has recently been applied in medicine, diagnostic imaging, and identifying and diagnosing abnormal regions(14,15). This study aimed to develop and evaluate an artificial intelligence–based algorithm that uses pre-RA IVUS images to automatically predict regions debulked by RA.

## Methods

### Study population

IVUS data from patients with de novo severely calcified coronary lesions who underwent IVUS-guided RA at five centers were consecutively collected from July 2016 to October 2021. IVUS imaging data were acquired using a VISICUBE IVUS imaging system with a 60-MHz mechanically rotating IVUS catheter (AltaView^TM^, Terumo, Tokyo, Japan). The criteria for lesion selection were as follows: (i) lesions located in the left anterior descending coronary artery (LAD), (ii) lesions with severe calcifications that were defined as radiopacities when observed without cardiac motion before contrast injection(12), (iii) IVUS examination performed before any intervention and immediately after RA, and (iv) the RA burr size was selected as 1.5 or 1.75 mm. The exclusion criteria were a diagnosis of acute coronary syndrome, in-stent restenosis, a branch lesion, left main narrowing >50%, and any balloon dilation before post-RA IVUS. The protocol for the retrospective data analysis was approved by our Institutional Review Board (2021252). The need for informed consent was waived because IVUS images were acquired during daily clinical practice from patients who consented to the comprehensive research use of their data. All the patients were guaranteed the opportunity to opt out of the study.

### RA and IVUS procedures

RA was performed under local anesthesia via the radial or femoral approach using a 6 or 7-Fr guiding catheter. After crossing the lesion with a 0.014-inch conventional guidewire, an IVUS image was obtained before any coronary intervention. The IVUS catheter was inserted into each coronary artery as distally as possible, and an imaging run was performed back to the aorto-ostial junction using an automated transducer pullback system. The decision to perform RA at each center was based on a clinical expert consensus document(8). Following the IVUS examination, the 0.014-inch conventional guidewire was exchanged with a 0.009-inch RotaWire floppy or RotaWire extra support guidewire (Boston Scientific, Marlborough, MA, USA) using a microcatheter. Each operator chose the guidewire and burr size for RA based on same document above(8). After the burr passed the lesion, it was removed using the Dynaglide mode. IVUS examinations were repeated immediately after RA in a manner similar to the pre-RA IVUS in all cases.

### IVUS segmentation

Preprocedural IVUS images were reviewed carefully to identify the cross-section with the minimum lumen area (MLA). IVUS analyses were conducted at the MLA at 0.5-mm intervals from sites 10 mm proximal to 10 mm distal to the MLA site. Cross-sections with poor-quality IVUS images were excluded from the study. The lumen contour was manually traced in every image frame, following a previously reported standard protocol(16,17) (Figure 1).

**Figure 1.**
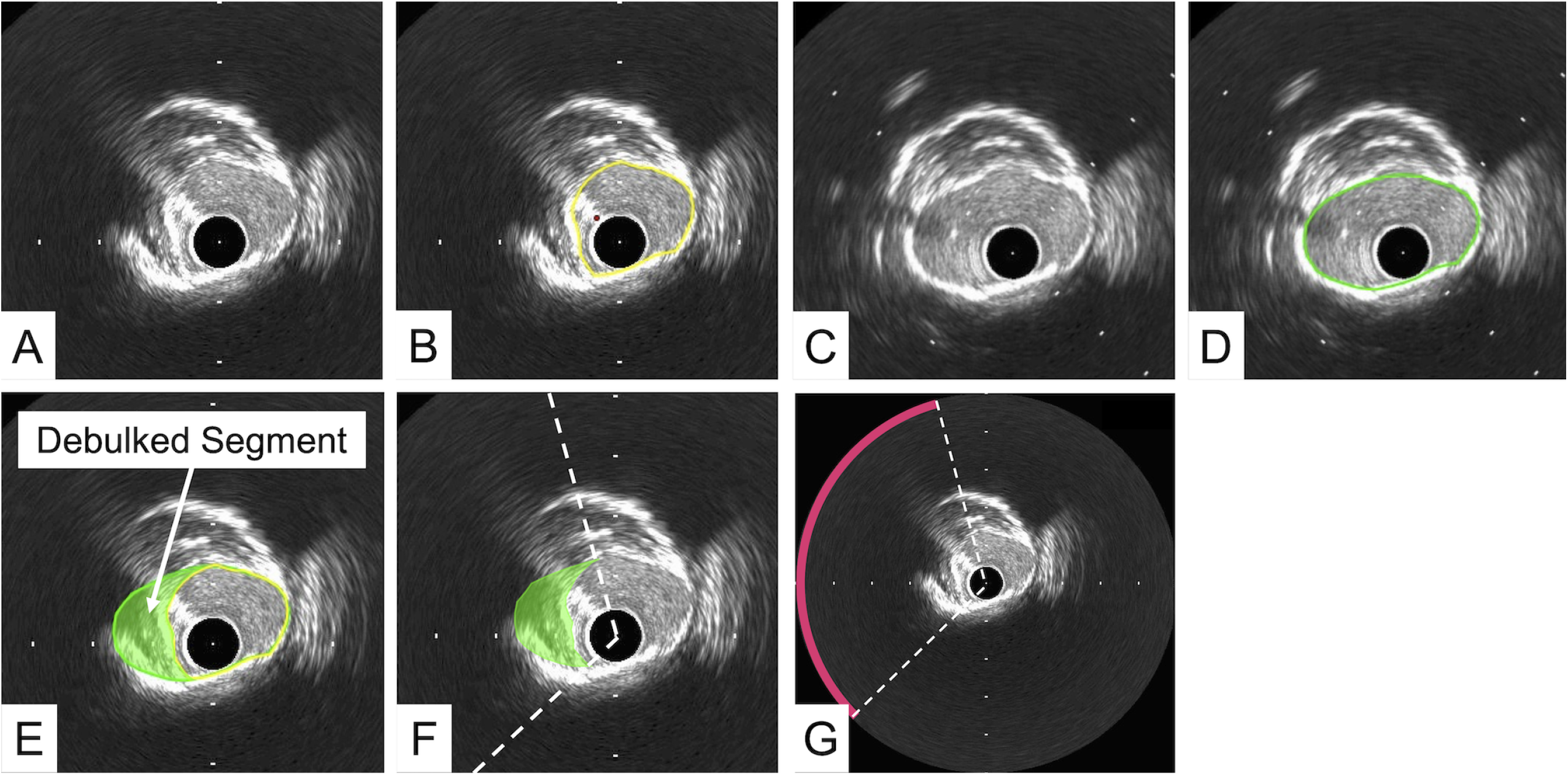
Analysis of IVUS images and creating ground truth labelling. (A) A pre-RA IVUS image. (B) Lumen contour on image A drawn by dedicated software. (C) A post-RA IVUS image matched with image A. (D) Lumen contour on image C. (E) A merged image of pre-RA IVUS image B and post-RA IVUS image D by the software. A debulked segment is identified by calculating the post-RA lumen area minus the pre-RA lumen area (green area). (F) The orientation of the debulked segments on the merged images. (G) The ground truth image shows the debulked segments’ orientation on the IVUS image’s outer circle. IVUS: intravascular ultrasound; RA: rotational atherectomy

Serial IVUS images taken before and immediately after RA were reviewed in parallel on the screen. The analyzed cross-sections were co-registered frame-by-frame based on the distance from landmarks, such as side branches or the site of calcification, following the same method of previous IVUS studies(12,18). The spatial axes of the two matched IVUS cross-sectional images were aligned and merged using dedicated software (ImageJ, NIH, Bethesda, MD, USA). The cross-section was considered to have a debulked region when the lumen area increased by >5% after RA, and the region with an enlarged lumen contained sharp traces of burr debulking on the post-RA IVUS image(12).

### Ground truth labelling

The orientations of the debulked segments identified in the merged images are marked on the outer circle of each IVUS image. Ground truth images were created by merging the same marks into the pre-IVUS images (Figure 1). Image creation was performed by a cardiologist using the software mentioned above.

### Model development

In our quest to further the capabilities of coronary artery imaging, we aimed to develop a deep-learning model capable of using IVUS images of coronary arteries to predict regions of excavation by the RA. The unique circular structure of the coronary arteries was utilized in our design. To achieve this, we partitioned the structure into 36 orientations, segmented every 10°. For each orientation, our model predicted whether a specific site would be debulked.

Another key feature of our model design is the continuity of the coronary arteries. Toward this end, the prediction for any slice *n* was obtained by incorporating images from slice *n*, one slice before (slice *n*−1), and one slice after (slice *n*+1), spaced 5 mm apart. We used three images to predict the excavation site for slice *n* more accurately. From an implementation perspective, our architecture connects 36 fully connected layers, each corresponding to one of the 36 orientations segmented every 10°, to a single feature extractor.

The model was trained such that the sum of the 36 individual loss values minimized the total loss. For our feature extractor, two architectures, ResNet [Deep Residual Learning for Image Recognition] and ConVNeXt [A ConvNet for the 2020s], were explored. Both employed pre-trained models from ImageNet and underwent fine-tuning. The Adam loss function was chosen for its optimization properties; data augmentation was implemented using RandAugment [Randaugment: Practical automated data augmentation with a reduced search space]. The model that achieved the smallest loss function value on the validation dataset (within 100 epochs) was selected as the best-performing model. The entire model was trained using the PyTorch framework [PyTorch: An Imperative Style, High-Performance Deep Learning Library]. Although our model was built from the ground up, it was inspired by and modified from the original source code available online (https://github.com/Medical-AI-Lab/Nervus) [Nervus: A Comprehensive DL Classification, Regression, and Prognostication Tool for both Medical Image and Clinical Data Analysis]. Specifically, the classifier from this codebase was adapted to accommodate the 36 orientations, which formed the foundation for our current model. A detailed model is shown in Figure 2; the source code is available online.

**Figure 2.**
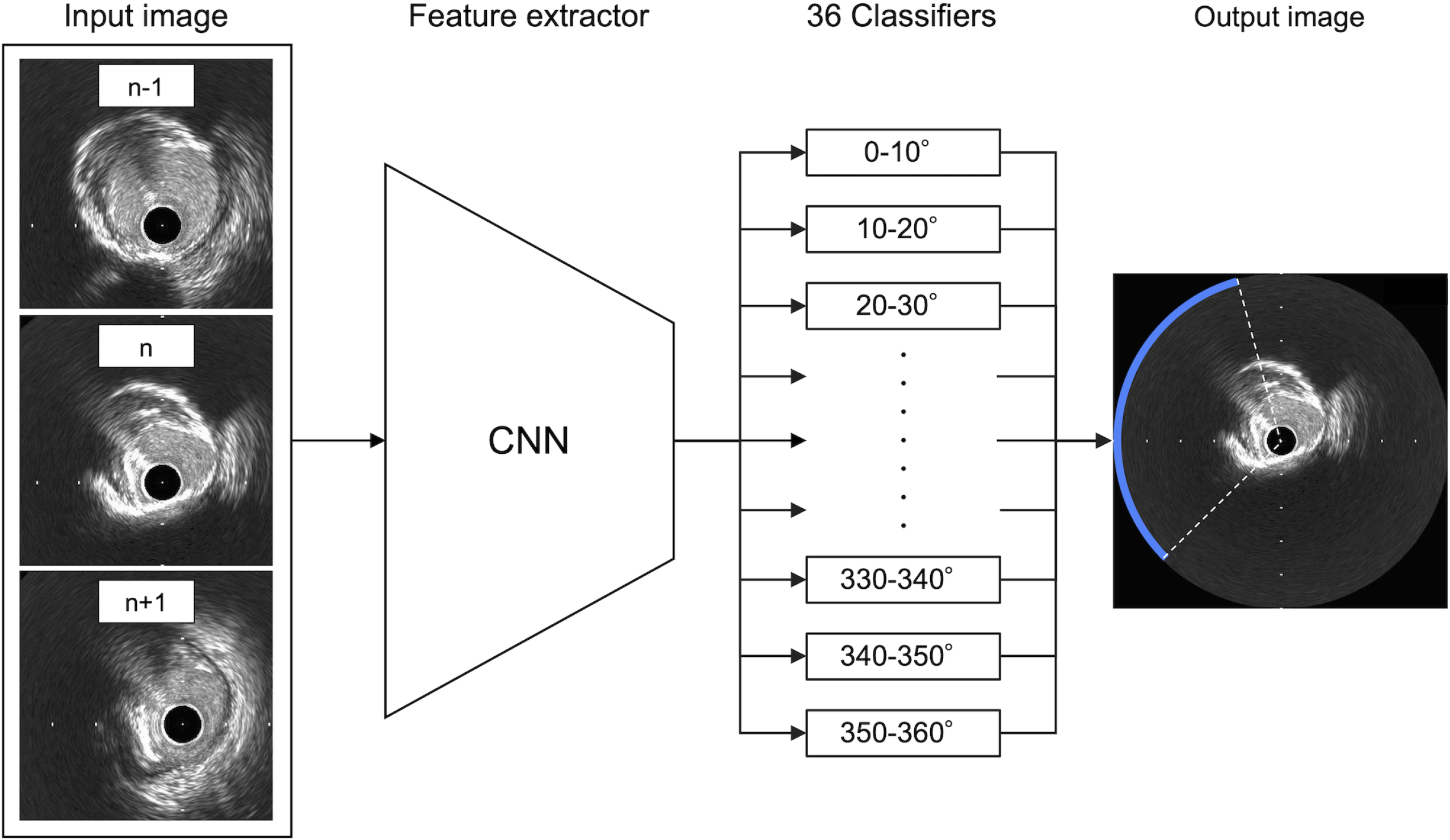
Overview of model development. In our model, the image of slice *n* and the images before (slice *n*−1) and after (slice *n*+1) are incorporated at 5-mm intervals. Using three input images, image information in the long-axis direction of the blood vessel is obtained, expecting to improve the prediction accuracy. ResNet is the base architecture for the proposed model because of its successful performance. Our architecture connects one feature extractor to 36 fully connected parallel layers (each layer corresponding to one of the 36 directions divided by 10°), which serve as classifiers. The 36 results and images of slice *n* are combined, and the deep-learning algorithm is coded so that they can be outputted.

### Model test

The performance of the best-performing model was assessed on the validation and test datasets. Sensitivity, specificity, positive predictive value (PPV), negative predictive value (NPV), and accuracy were used as primary metrics for evaluation. These metrics were presented as averages for each degree and image.

### Statistical analysis

All statistical analyses of the model tests were performed using R version 4.0.0. For statistical inferences, we adopted a two-sided significance level of 5% and relied on the Clopper–Pearson method.

## Results

### Data

The study analyzed 2460 cross-sections from 60 LAD lesions in 60 patients. Because the cross-sectional images were of low quality or cross-sectional images from slice n+1 (5 mm distal from the MLA site) and/or slice n-1 (5 mm proximal from the MLA site) were missing, 354 images were excluded. Consequently, 2106 cross-sections were segmented and labeled as the ground truth. Among these cross-sections, 1416 (67%) contained debulked regions; the remaining 690 (33%) did not. The images were divided into training (1649), validation (226), and test (231) datasets.

### Model development

Upon evaluating the performances of various architectures, we found that ResNet had superior accuracy and efficiency. Its sophisticated design and deep layers demonstrate a distinct advantage, making it the leading choice for our artificial intelligence model for predicting debulked regions from pre-RA IVUS images of the coronary arteries. During this period, the model with the lowest loss in the validation data was considered the best-performing model. The model was developed using the training and validation datasets.

### Diagnostic performance of our artificial intelligence model

The best-performing model was evaluated using test datasets. In each cross-sectional analysis, our artificial intelligence model achieved an average sensitivity, specificity, PPV, NPV, and accuracy of 81%, 72%, 46%, 90%, and 75%, respectively. Table 1 shows the 10°-wise performances for predicting the region debulked by RA. In the 10°-wise analysis, no significant difference was found in diagnostic accuracy among the individual 10° regions. A representative image of the proposed model is shown in Figure 3.

**Figure 3.**
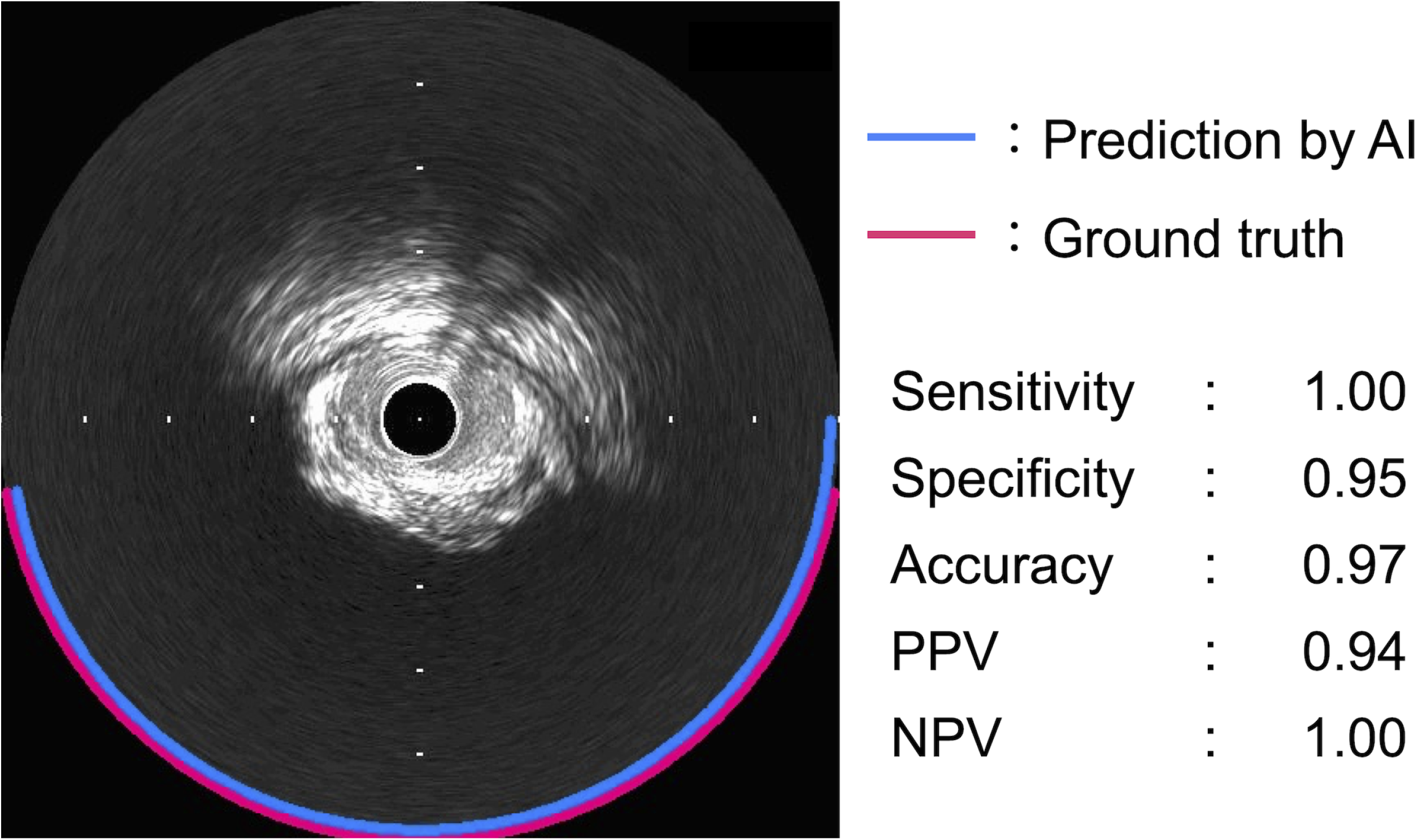
Representative image of model test. The diagnostic performance of our model is evaluated based on the agreement rate between the debulking angle predicted by the model and the debulking angle of the ground truth. It is evaluated using 457 datasets allocated as validation and testing datasets. Here, we present a representative case with acceptable test results.

**Table 1.** The cross-sectional and the 10°-wise performances for predicting the region debulked by RA.

|  | Sensitivity | Specificity | Accuracy | PPV | NPV |
| --- | --- | --- | --- | --- | --- |
| Cross-sectional level | 81 | 72 | 75 | 46 | 90 |
| 10°-wise level |  |  |  |  |  |
| 0 - 10° | 58 | 84 | 79 | 47 | 89 |
| 10 - 20° | 80 | 85 | 80 | 52 | 89 |
| 20 - 30° | 62 | 84 | 79 | 53 | 88 |
| 30 - 40° | 80 | 68 | 71 | 46 | 91 |
| 40 - 50° | 90 | 63 | 70 | 46 | 95 |
| 50 - 60° | 88 | 68 | 73 | 51 | 93 |
| 60 - 70° | 89 | 54 | 65 | 47 | 91 |
| 70 - 80° | 83 | 59 | 67 | 51 | 87 |
| 80 - 90° | 86 | 58 | 67 | 50 | 89 |
| 90 - 100° | 85 | 65 | 72 | 58 | 88 |
| 100 - 110° | 89 | 49 | 64 | 50 | 89 |
| 110 - 120° | 86 | 56 | 67 | 53 | 87 |
| 120 - 130° | 87 | 55 | 66 | 51 | 88 |
| 130 - 140° | 82 | 60 | 68 | 53 | 86 |
| 140 - 150° | 85 | 64 | 72 | 59 | 88 |
| 150 - 160° | 88 | 68 | 75 | 62 | 91 |
| 160 - 170° | 92 | 61 | 72 | 57 | 93 |
| 170 - 180° | 93 | 66 | 75 | 58 | 95 |
| 180 - 190° | 95 | 72 | 80 | 64 | 96 |
| 190 - 200° | 87 | 77 | 80 | 63 | 93 |
| 200 - 210° | 66 | 82 | 77 | 62 | 85 |
| 210 - 220° | 68 | 77 | 74 | 55 | 85 |
| 220 - 230° | 62 | 77 | 73 | 50 | 84 |
| 230 - 240° | 72 | 80 | 78 | 56 | 89 |
| 240 - 250° | 83 | 83 | 83 | 59 | 94 |
| 250 - 260° | 86 | 84 | 84 | 60 | 96 |
| 260 - 270° | 86 | 88 | 86 | 60 | 96 |
| 270 - 280° | 88 | 80 | 81 | 50 | 97 |
| 280 - 290° | 83 | 87 | 87 | 59 | 96 |
| 290 - 300° | 79 | 84 | 83 | 49 | 95 |
| 300 - 310° | 77 | 81 | 81 | 45 | 95 |
| 310 - 320° | 79 | 80 | 80 | 45 | 95 |
| 320 - 330° | 63 | 79 | 77 | 38 | 92 |
| 330 - 340° | 85 | 78 | 79 | 45 | 96 |
| 340 - 350° | 83 | 81 | 79 | 47 | 93 |
| 350 - 360° | 58 | 84 | 79 | 47 | 89 |

RA: rotational atherectomy; PPV: positive predictive value; NPV: negative predictive value.

## Discussion

The artificial intelligence–based algorithm developed in this study shows promise for using pre-RA IVUS images to predict which regions of the coronary plaque would be debulked by RA. The main results of this study were 1) we developed a deep-learning algorithm capable of automatically predicting the orientation of the regions debulked by RA based on pre-RA IVUS images, and 2) the diagnostic accuracy of the developed artificial intelligence model was approximately 80%.

Because of an aging population with an increasing burden of cardiovascular risk factors, complex calcified coronary lesions are increasingly encountered in daily clinical practice. Intravascular imaging is currently recommended as an auxiliary device for safely performing RA in the guidelines(8). Previous studies on predicting RA debulking regions using intravascular imaging have shown that the location of the guidewire and imaging catheter can predict coronary plaque debulking regions; the vessel wall closer to the guidewire tends to be more debulked than the vessel wall closer to the imaging catheter(11,12). However, the diagnostic accuracy was low at 67% in studies using IVUS(12) and 48% in studies using optical coherence tomography(11). These rates are insufficient to prevent catastrophic events such as coronary perforation in routine daily clinical practice. Furthermore, these diagnostic accuracies were calculated based on the analysis of cross-sectional images with debulked regions only, not on analyzing all cross-sections imaged by IVUS, as in this study. When all cross-sectional images obtained by IVUS were included in the analysis of a previous study, the concordance rate between the guidewire position and the debulked region was only 42%(12). From this perspective, the 80% diagnostic accuracy for predicting the debulked region with RA from pre-RA IVUS achieved by the artificial intelligence model developed in this study significantly exceeded that of human predictions. To our knowledge, this study is the first to demonstrate that an artificial intelligence model can use pre-RA IVUS images to accurately predict which parts of a plaque would be debulked by RA.

Recent technological advances have facilitated the application of artificial intelligence models for medical image analysis. In cardiovascular disease, the application of artificial intelligence to electrocardiography(19,20) and chest X-rays(15,21) has enhanced the interpretation of images and decision-making in treatment strategies. Only a few studies have applied artificial intelligence to clinical practice for intracoronary imaging, particularly intravascular ultrasound imaging. Cho et al.(22) reported an artificial intelligence model that could automatically classify IVUS frames (with or without calcification and ultrasound attenuation) with a high overall accuracy of 90%. The extent of ultrasound attenuation and calcification in the ROI segment can be quantified accurately within a few seconds of inputting the IVUS image into the model. In addition, another study reported the high-accuracy detection of stent underexpansion sites from IVUS images after stent implantation using an artificial intelligence model(23). Unlike transthoracic echocardiographic images, the quality of the acquired IVUS images is independent of the operator’s skill and experience. The ability to predict the region debulked by the RA independent of experience is advantageous for less experienced PCI operators. This finding is particularly relevant because both the number of RA cases per operator and per year are inversely associated with the incidence of RA-related complications(9,24). In addition, the model can be utilized by PCI operators at any time, which is especially valuable in areas where experienced PCI operators are unavailable or during nighttime emergencies when the medical staff is limited. From this perspective, the artificial intelligence model is advantaged over an intuitive evaluation based on the experience of the PCI operator performing the RA procedure.

### Limitation

This study had some limitations. First, our artificial intelligence model was developed and validated only for the LAD and did not support the right coronary and left circumflex arteries. Second, observing lesions using IVUS before RA is essential for applying our results in routine clinical practice. However, the IVUS imaging catheter may not be advanced beyond severely calcified lesions that require RA. Third, the model was developed from IVUS images of cases using RA-burr sizes of 1.5 and 1.75 mm; the diagnostic accuracy when using other burr sizes is unknown. Fourth, the wire used during IVUS differed from that used during RA.

## Conclusions

The artificial intelligence–based algorithm can use information from pre-RA IVUS images to accurately predict regions debulked by RA. These results are highly promising. The proposed method will assist interventional cardiologists in determining the treatment strategies for severely calcified coronary lesions.

## Clinical Perspectives

### Clinical Competencies

Although rotational atherectomy is a useful technique for severely calcified lesions, patients undergoing RA show a greater incidence of catastrophic complications, such as coronary perforation. Therefore, we have empirically predicted the segment where the atherectomy burr debulked from intravascular ultrasound images, but the diagnostic accuracy of human experience-based prediction is low.

### Translational Outlook

Although the diagnostic accuracy was high, the positive predictive value was insufficient. This model needs to be tested prospectively in real clinical cases to investigate its impact on clinical practice in the field of coronary intervention.

### Abbreviations

IVUS: intravascular ultrasound
LAD: left anterior descending coronary artery
MLA: minimum lumen area
NPV: negative predictive value
PCI: percutaneous coronary intervention
PPV: positive predictive value
RA: rotational atherectomy

## Data Availability

Our model was inspired by and modified from the original source code available online [Nervus: A Comprehensive DL Classification, Regression, and Prognostication Tool for both Medical Image and Clinical Data Analysis].

https://github.com/Medical-AI-Lab/Nervus

## Acknowledgments

The authors thank the catheterization laboratory staff and the Clinical Research Center staff for their excellent support of the study.

## Notes

### Competing Interest Statement

The authors have declared no competing interest.

### Clinical Trial

The protocol for the retrospective data analysis was approved by Institutional Review Board of Kansai Medical University (2021252)

### Funding Statement

This study was supported by the Research Grant D2 from Kansai Medical University, Hirakata, Japan, and a grant from an Academic Support Award: Japanese Association of Cardiovascular Intervention and Therapeutics, Tokyo, Japan.

